# Clonal Hematopoiesis Associates with Prevalent and Incident Cardiometabolic Disease in High-Risk Individuals

**DOI:** 10.1101/2025.01.14.25320566

**Authors:** Jessica A. Regan, Lydia Coulter Kwee, Navid A. Nafissi, Alexander G. Bick, William E. Kraus, Pradeep Natarajan, Sidd Jaiswal, Svati H. Shah

## Abstract

**Background:** Clonal hematopoiesis of indeterminate potential (CHIP) is the age-related presence of expanded somatic clones secondary to leukemogenic driver mutations and is associated with cardiovascular (CV) disease and mortality. We sought to evaluate relationships between CHIP with cardiometabolic diseases and incident outcomes in high-risk individuals.

**Methods:** CHIP genotyping was performed in 8469 individuals referred for cardiac catheterization at Duke University (CATHGEN study) to identify variants present at a variant allele fraction (VAF) ≥2%. Associations were tested among any CHIP variant, large CHIP clones (VAF ≥10%) and individual CHIP genes with prevalent cardiometabolic traits. Cox proportional hazard models tested CHIP associations with time-to-overall mortality and Fine-Gray analyses tested CHIP associations with incident cardiovascular outcomes.

**Results:** We identified 463 CHIP variants in 427 individuals (5.0%) of which 268 (3.2%) harbored large CHIP clones. CHIP and large CHIP were associated with lower odds of obesity (OR 0.79 [95% CI 0.65-0.98], p=0.03; OR 0.76 [95% CI 0.57-0.99], p=0.04, respectively). CHIP was associated with prevalent HF (OR 1.25 [95% CI 1.01 - 1.55], p=0.04; especially for non-*DNMT3A* CHIP (OR 1.38 [95% CI 1.04-1.82], p=0.02). CHIP was also associated with incident events: Non-*DNMT3A* CHIP was associated with increased risk of time-to-HF hospitalization (HR 1.29 [95% CI 1.02-1.63], p=0.03).

**Conclusions:** In high-risk individuals referred for cardiac catheterization, large CHIP and non-*DNTM3A* CHIP were associated with obesity, prevalent HF, incident CV events. These findings strengthen the importance of CHIP as a biomarker for CV disease and highlight the contributing risk of large CHIP clones and non-*DNMT3A* CHIP variants.

**Condensed Abstract:** CHIP, the presence of somatic expanded mutations in myeloid driver genes in hematopoietic cells, is an emerging CVD biomarker. Using whole exome sequencing of peripheral blood derived DNA from participants in the CATHGEN cohort, we identified significant associations with obesity, prevalent HF, incident mortality, HF hospitalization and AF after adjusting for established clinical risk factors. These findings add strength to the growing literature of CHIP as a CVD biomarker, emphasizing large CHIP and non-*DNMT3A* CHIP variants for driving risk. Future studies should aim to further elucidate gene-specific risk and the inflammatory and metabolic mechanisms possibly mediating these relationships.

**Clinical Perspective:** *What Is New?:*

- In a cohort with high prevalence of CAD, CHIP is inversely associated with obesity and associated with higher odds of prevalent HF and subsequent mortality, even after adjustment for relevant clinical comorbidities. Risk of incident events of mortality, HF hospitalization and AF were driven by large CHIP variants (VAF≥10%) and CHIP variants in genes other than *DNMT3A*.

*What are the Clinical Implications?:*

- Though more research is needed, as the evidence around the risk associated with specific CHIP variants continues to grow, clinicians should be prepared to provide gene-specific counseling for cardiometabolic disease risk.

## Introduction

Genetic epidemiology studies have identified hundreds of variants associated with cardiovascular disease (CVD); however, these variants typically have small effect sizes and incompletely predict CVD risk^1^. While work has focused on germline mutations, which occur with formation of the embryo and are static over the lifetime, somatic mutations have increasingly been shown to contribute to CVD. Inherent in cancers, somatic mutations have been identified in diverse non-tumor tissues including in the atria of patients with atrial fibrillation (AF)^2^.

Clonal hematopoiesis (CH) is the age-related expansion of somatic clones in hematopoietic stem cells (HSCs). CH of indeterminate potential (CHIP) can be detected in peripheral blood DNA and is defined as having a clone size (variant allele fraction [VAF]) ≥2% in genes associated with the development of myeloid malignancies and myelodysplastic syndrome (MDS)^3–5^. The most frequently mutated CHIP genes are *DNMT3A*, *TET2* and *ASXL1*, where mutant cells have a competitive proliferative advantage over native HSCs. Germline contributors to acquisition of CHIP have been identified^6–8^, and smoking has been associated with increased odds of *ASXL1*-mutated CHIP^6^.

The presence of CHIP is associated with increased risk of overall mortality, which appears to be driven by CVD events more so than by malignancy-related mortality. CHIP is associated with increased risk of incident coronary artery disease (CAD) and accelerated atherosclerosis, with heightened levels of inflammation in both clinical and pre-clinical models^9,10^. Large CHIP clones (VAF >10%) in *DNMT3A* and *TET2* contribute greater risk for incident CVD^11^. CH is also associated with incident heart failure (HF)^8,12,13^ including incident heart failure with preserved ejection fraction (HFpEF)^14,15^, as well as AF^8,16–18^. Variants in particular CHIP genes, have been identified as drivers of phenotype-specific risk such as *TP53* and atherosclerosis^19^ and *TET2* and HF^13–15^.

Knowledge gaps remain in our understanding of CHIP in patients with high cardiometabolic risk and existing cardiovascular disease. Although *DNMT3A* is the most commonly mutated CHIP gene, these variants may not be the strongest drivers of cardiometabolic risk^13^. Given the evolving CVD phenotypic associations, we sought to identify novel findings of CHIP, large CHIP and mutations in genes other than *DNMT3A* (non-*DNMT3A*) and cardiometabolic traits and to assess further validation for CVD events, HF and AF in a high-risk population. Here we leverage whole exome sequencing (WES) to explore the role of CHIP as an emerging biomarker for intermediate cardiometabolic risk factors and prevalent and incident CVD in a medically complex, but clinically relevant cohort of patients referred for cardiac catheterization.

## Methods

### Study Population

The CATHGEN biorepository is comprised of 9334 individuals who underwent cardiac catheterization at Duke University Medical Center (Durham, NC) between January 2001 and December 2010^20^. Femoral arterial blood samples were collected at the time of catheterization and whole blood was stored in ethylenediaminetetraacetic acid (EDTA) tubes. All study participants gave written informed consent for participation and use of their stored biospecimens for future use. The study was approved by the Duke University Institutional Review Board.

Demographics and comorbidities were collected through medical record review at study enrollment, and yearly follow-up was conducted for events and vital status through 2020. These data were supplemented with electronic health records, including International Classification of Diseases, Ninth Revision (ICD-9) and Tenth Revision (ICD-10) codes. Patients with a diagnosis of hematologic malignancy prior to or within 6 months of study enrollment were excluded for this study (n=99, **Supplemental Table 1,** “Prevalent Hematologic Malignancy” phenotype).

### Whole Exome Sequencing and Somatic Variant Calling

WES was performed by Regeneron Genetics on DNA extracted from EDTA whole blood using the HiSeq 2500 platform (Illumina, San Diego, California). Following genomic sequence alignment and quality control, the resultant BAM files were used for somatic variant calling using the GATK^21^ Mutect2 pipeline on the Terra platform^22,23^. A panel of normals (PON) created from 40 young, healthy participants was used to eliminate sequencing artifacts. Functional annotation was performed for identified somatic variants and output into a variant call format (VCF) file. VCF files were parsed to regions of interest in 68 previously described CHIP genes and filtered to specific single-nucleotide variants (SNVs) and indels (**Supplemental Table 2**).

CHIP was defined as presence of a variant at a VAF ≥2% and large CHIP clones at a VAF >10%. An experienced hematopathologist performed manual curation and review to refine the final variant call set. Additional details on sequencing and variant calling details can be found in the **Supplemental Methods**. After WES quality control, 8469 participants had high quality somatic variant calls and were included for analyses. For participants with variants in multiple CHIP genes identified, analyses are categorized by the largest CHIP clone.

### Clinical Data and Outcomes

Baseline cardiometabolic comorbidities were assessed by the enrolling physician and medical record review. Obesity was defined as BMI ≥30kg/m^2^. Prevalent CAD was defined as a prior history of MI, coronary artery bypass surgery, coronary percutaneous intervention, or cardiac catheterization with the presence of more than one major epicardial coronary vessel with 50-74% stenosis or one vessel with ≥75% stenosis. Prevalent CAD, HF and AF diagnoses were supplemented by ICD-9 and ICD-10 codes within six months or prior to index catheterization. All-cause mortality was determined using the Social Security Death Index (SSDI), National Death Index (NDI) and follow-up calls through the Duke Information System for Cardiovascular Care; cause of death was available on a subset of patients. ICD-9 and ICD-10 codes for emergency room visits or hospitalizations between 2001 and 2020 were used to define incident outcomes: MI and HF hospitalization (**Supplemental Table 1**, “Myocardial Infarction” and “Heart Failure / Cardiomyopathy” phenotype, respectively). Patients with MI within 30 days of their HF outcome were excluded to avoid new diagnoses of acute heart failure secondary to MI^24^. Incident AF was defined by ICD-9 or ICD-10 codes (**Supplemental Table 1**, “Atrial Fibrillation” phenotype).

### GRACE Score

Although not all CATHGEN participants presented with acute coronary syndrome, given the high prevalence of baseline CAD (64.6%) in a cardiac catheterization cohort we used the Global Registry of Acute Coronary Events (GRACE) score to determine the prognostic significance of CHIP for overall mortality prediction in addition to a validated clinical score^25^ (**Supplemental Methods**).

### Statistical Analyses

A summary of the analytic approach is shown in **Supplemental Table 3**. Associations of age with CHIP and clone size were tested using two sample t-test. Associations of CHIP, large CHIP and specific CHIP genes and obesity were tested using logistic regression in univariate and multivariate models. Across all multivariate analyses the basic covariates adjusted for were: age, genetic ancestry, sex, smoking.

Analyses of CHIP and other prevalent cardiometabolic factors CAD, HF and AF were additionally adjusted for diabetes, BMI, hypertension and hyperlipidemia. Sensitivity models for prevalent CAD included statin therapy (available in 61.2% of participants). HF and AF models were additionally adjusted for prevalent CAD and history of malignancy given the known associations of chemotherapy exposure with both CHIP and cardiomyopathy^24^, and AF was also adjusted for prevalent HF. Stratified sensitivity analyses tested the association between HF phenotypes and CHIP: HFpEF (LVEF≥50%) and HF with reduced ejection fraction (HFrEF, LVEF<50%) using ANOVA and chi-square tests to test CHIP proportions in these groups compared to participants without HF^26^. These HF phenotype sensitivity analyses excluded participants with a history of congenital heart disease, valvular disease, heart transplant or end-stage renal disease requiring dialysis.

Associations of CHIP and time-to-overall mortality were tested using Cox proportional hazard models and compared to the GRACE score. Fine-Gray competing risk regression models were used to test associations for incident outcomes with overall mortality as a competing risk: composite of MI or CV death, HF hospitalization and AF. Incident outcome analyses were adjusted for the same covariates as above including prevalent CAD, HF and AF. Given the long length of follow-up sensitivity analyses also truncated follow-up duration at ten years.

For each clinical outcome of interest, analyses were performed to test associations with the presence of any CHIP variant and large CHIP (VAF>10%). To determine gene-specific risk, the most frequently mutated CHIP genes: *DNMT3A*, *TET2*, *ASXL1* and non-*DNMT3A* CHIP were also tested, requiring a minimum of ten cases or events for gene level analyses. Significance was considered at p<0.05. Analyses were performed in R version 4.4.2.

## Results

### Baseline Clinical Characteristics and CHIP Variants Identified

Baseline participant characteristics are shown in **Table 1**. We identified an overall prevalence of CHIP of 5.0% with 463 CHIP variants in 427 unique participants, including 210 *DNMT3A*, 100 *TET2*, and 45 *ASXL1* variants (**Supplemental Table 3, Supplemental Figure 1a**). The median VAF across all CHIP variants was 13.2% (IQR 7.9-22.5%), including 289 large CHIP clones in 268 individuals (**Supplemental Figure 1b**). As expected, participants with CHIP were older (mean 69.5±10.3 years) than those without CHIP (60.8±12.0 years, p<2×10^-16^, **Supplemental Figure 2**). Individuals with large CHIP clones were on average 2.9 years older (mean 70.6±9.7 years) than those with small CHIP clones (VAF<10%, 67.7±11.0 years, p=0.006).

**Table 1:**
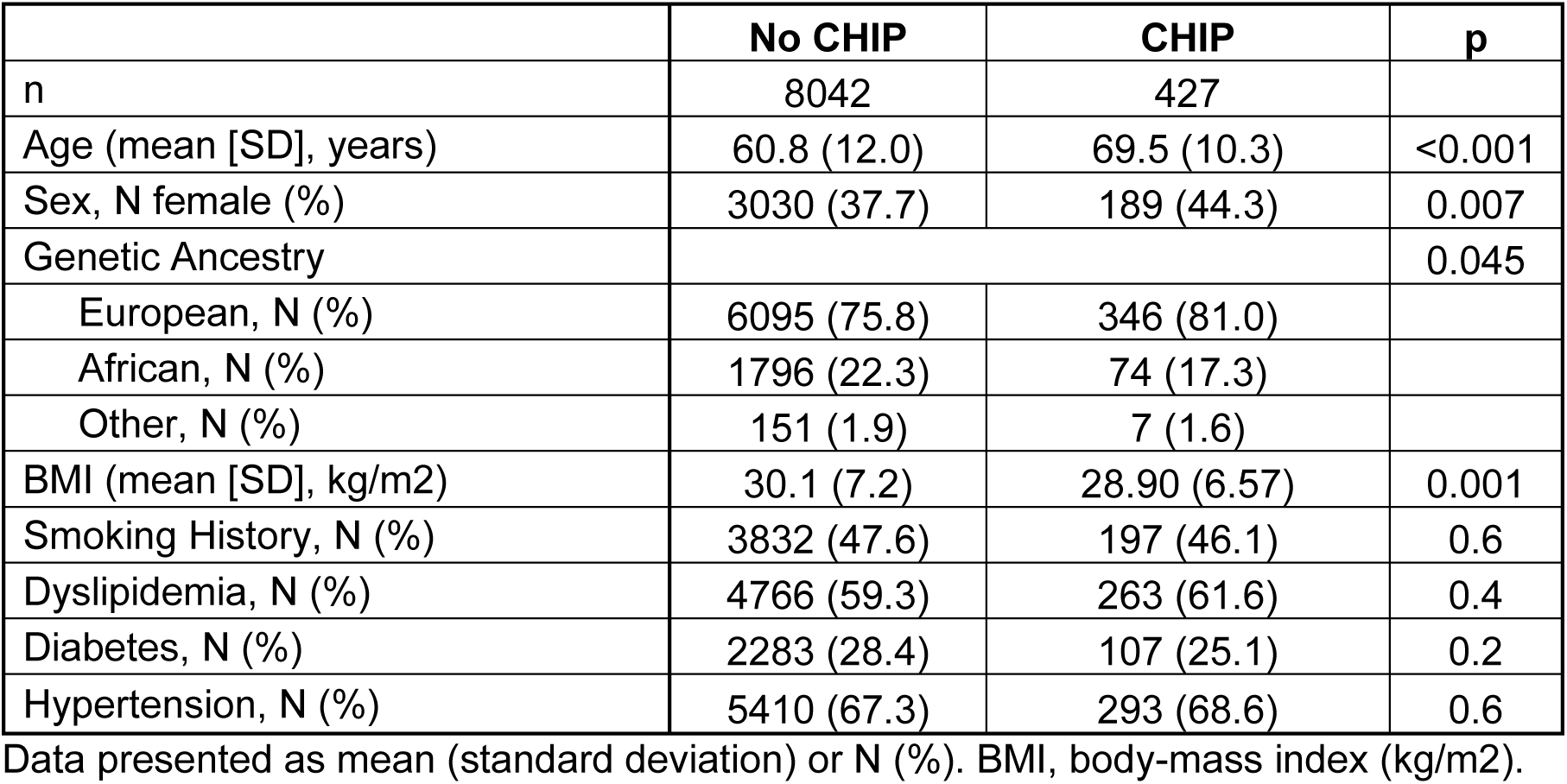
Baseline Participants Characteristics Stratified by CHIP Status.

**Table 2.** Summary of Significant Associations of CHIP with Prevalent and Incident Cardiometabolic Disease in CATHGEN.

| Gene | Prevalent Outcomes |  |  |  | Incident Outcomes |  |  |  |
| --- | --- | --- | --- | --- | --- | --- | --- | --- |
|  | Obesity | CAD | HF | AF | Mortality | MI or CV death | HF Hospitalization | AF |
| CHIP | ↓ / ↓ | - | ↑ / ↑ | ↑ / - | ↑ / - | - | - | - |
| Large CHIP | ↓ / ↓ | - | ↑ / - | ↑ / - | ↑ / ↑ | ↑ / - | - | - |
| non-*DNMT3A* | ↓ / - | - | ↑ / ↑ | ↑ / - | ↑ / ↑ | ↑ / - | ↑ / ↑ | ↑ / - |
| *DNMT3A* | ↓ / - | - | - | ↑ / - | ↑ / - | - | ↓ / ↓ | - |
| *TET2* | - | - | - | ↑ / - | ↑ / - | - | - | - |
| *ASXL1* | ↓ / - | ↑ / - | ↑ / - | ↑ / - | ↑ / - | ↑ / - | ↑ / - | ↑ / ↑ |

Summary of significant associations for prevalent and incident outcomes analyzed. The first arrow indicates the direction of significant univariate associations, and the second arrow indicates significant multivariate associations; "-", indicates no significant association. CAD, coronary artery disease; HF, heart failure; AF, atrial fibrillation; MI, myocardial infarction; CV, cardiovascular.

Participants with CHIP were more often female (44.3%) than those without CHIP (37.7%, p=0.007). *DNMT3A* CHIP carriers were mostly female (50.5%), whereas *TET2* and *ASXL1* CHIP carriers were predominantly male (55.8% and 81.3%, respectively). Participants with CHIP had a higher percentage of European ancestry (81.0%) than those without CHIP (75.8%, p=0.045). Participants with CHIP also had a lower BMI (28.9±6.6 kg/m^2^) than those without CHIP (30.1±7.2 kg/m^2^ p=0.001). There were no differences in history of smoking, dyslipidemia, diabetes or hypertension for overall CHIP, though 72.1% of *ASXL1* CHIP carriers had a history of smoking.

Thirty-two individuals had greater than one CHIP variant, five of which had two *TET2* variants, five had *TET2* and *DNMT3A* co-mutations and three had two *DNMT3A* variants (**Supplemental Figure 3**). Individuals with multiple CHIP variants were older (mean 72.5±8.2 years, p=0.045) and had greater history of MI (46.9% vs. 26.1%, p=0.02) and coronary artery bypass surgery (46.9% vs. 20.5%, p=0.001) than those with one CHIP variant.

### CHIP is Associated with Prevalent Cardiometabolic Disease

CHIP and large CHIP were inversely associated with obesity in univariate models and multivariate models (adjusted OR [aOR] CHIP 0.79 [95% CI 0.64-0.98], p=0.03; aOR large CHIP 0.76 [95% CI 0.57-0.99], p=0.04, **Supplemental Figure 4, Supplemental Table 5**). Non-*DNMT3A*, *DNMT3A* and *ASXL1* CHIP were inversely associated with obesity in univariate models, but the relationship was attenuated in multivariate models.

There were no significant associations with CHIP or large CHIP in univariate or multivariate analyses with prevalent CAD (**Supplemental Table 5**). *ASXL1* CHIP was associated with CAD in univariate models (OR 5.36 [95% CI 2.16-17.89], p=0.001), but was not significant in multivariate models (**Supplemental Table 5**). There were no significant relationships in sensitivity models adjusting for baseline statin therapy.

CHIP and large CHIP were associated with prevalent HF in univariate models, though only overall CHIP remained significant in multivariate analyses (aOR 1.25 [95% CI 1.01 - 1.55], p=0.04, **Figure 1, Supplemental Table 5**). Gene level analysis revealed that non-*DNMT3A,* and *ASXL1* CHIP carriers had a higher odds of prevalent HF in univariate analyses, only non-*DNMT3A* remained significant in multivariate analyses (aOR 1.38 [95% CI 1.04-1.82], p=0.02). Sensitivity analyses of CHIP and HF phenotypes suggested the highest prevalence of CHIP in HFrEF (6.1%) vs. HFpEF (5.0%) vs. No HF (4.5%; ANOVA p=0.04, chi-squared p=0.04 for HFrEF vs. No HF, **Supplemental Figure 5**).

**Figure 1.**
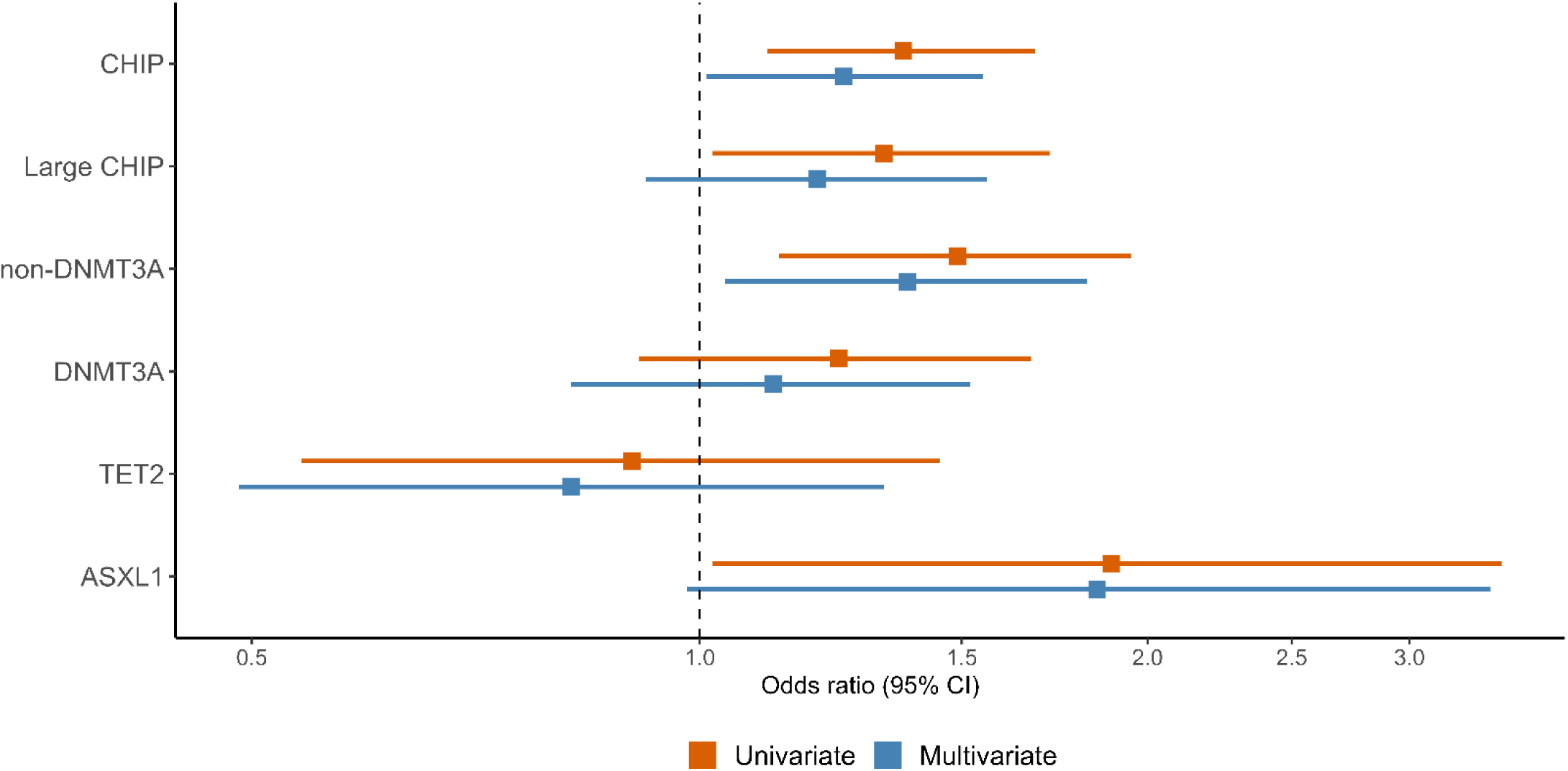
CHIP and non-*DNMT3A* CHIP Associate with Prevalent HF The presence of any CHIP variant and non*-DNMT3A* was associated with higher odds of prevalent HF in CATHGEN. Multivariate models are adjusted for age, sex, genetic ancestry, smoking, diabetes, body-mass index, hypertension, hyperlipidemia, prevalent coronary artery disease and history of malignancy.

CHIP and large CHIP were associated with higher odds of prevalent AF in univariate models (OR CHIP 1.72 [95% CI 1.37-2.14], p=1.8×10^-6^, OR large CHIP 1.75 [95% CI 1.32-2.30], 6.6×10^-5^), but the relationship was not significant in multivariate models (**Supplemental Table 6**). Similarly, non-*DNMT3A*, *DNMT3A*, *TET2* and *ASXL1* CHIP were associated with higher odds of prevalent AF in univariate, but not multivariate models.

### CHIP Associates with Overall Mortality in CATHGEN

The median follow-up time was 9.97 years (IQR 5.57-12.95 years) and there were 4197 total deaths. The presence of any CHIP variant and large CHIP were associated with time-to-death in univariate, but only large CHIP remained associated with time-to-death in multivariate models (aHR 1.17 [95% CI 1.01-1.36], p=0.04, c-statistic=0.675, **Supplemental Table 6, Supplemental Figure 6**). The presence of CHIP in non-*DNMT3A*, *DNMT3A*, *TET2* and *ASXL1* were strongly associated with time-to-death in univariate models, only non-*DNMT3A* remained associated in multivariate models (aHR 1.31 [95% CI 1.12-1.54], p=0.001, c-statistic=0.676, **Supplemental Table 6, Supplemental Figure 6**).

Given the high prevalence of baseline CAD in a cohort referred for catheterization we used the GRACE score to determine the prognostic value of CHIP in addition to a validated clinical score for predicting mortality. Mean GRACE Score was higher in participants with CHIP, non-*DNTM3A* and large CHIP (92.0±21.2, 93.1±21.9 and 94.5±20.7 points, respectively) versus those without CHIP (77.4±22.0 points). The GRACE Score was strongly associated with mortality (aHR 1.02 [95% CI 1.01-1.02], p<2.0×10^-16^, c-statistic=0.682), where an increase in HR is associated with the risk associated with every additional GRACE score point. The addition of either large CHIP or non-*DNMT3A* CHIP to the GRACE score did not improve the c-statistic. In sensitivity models truncated at ten years, non-*DNMT3A* CHIP remained associated with time-to-death, but large CHIP was not. There were no significant differences in the predictive capability of the GRACE score or addition of non-*DNMT3A* CHIP at ten years.

### CHIP Associates with Incident Cardiovascular Disease

Over a median follow-up of 9.50 years (IQR 5.02-12.74 years), a total of 2286 participants suffered a composite outcome of time-to-first incident MI or CV death. While overall CHIP was not associated with the composite CV outcome, large CHIP clones were associated in univariate models (HR 1.37 [95% CI 1.11-1.68], p=0.003) but this relationship was attenuated in multivariate models (aHR 1.21 [95% CI 1.01-1.44], p=0.056, **Supplemental Table 7**).

Similarly non-*DNMT3A* and *ASXL1* CHIP were associated with higher risk of the composite CV outcome in univariate models but were no longer significant in multivariate models (**Supplemental Table 7**).

Over a median follow-up of 8.08 years (IQR 3.21-12.00 years), there were 2541 participants with incident HF hospitalization. There were no significant associations for overall CHIP or large CHIP in univariate or multivariate models for incident HF hospitalization. Non-*DNMT3A* and *DNMT3A* CHIP were associated with incident HF hospitalization in both univariate and multivariate models, but with opposite direction of effect. Non-*DNMT3A* CHIP was associated with higher risk of HF hospitalization (aHR 1.29 [95% CI 1.02-1.63], p=0.03), whereas the presence of *DNMT3A* CHIP was associated with lower risk of incident HF hospitalization (aHR 0.65 [95% CI 0.48-0.88], p=0.005, **Supplemental Table 7, Figure 2**). Non-*DNMT3A* and *DNMT3A* CHIP remained associated with time-to-HF hospitalization in sensitivity models truncated at ten years.

**Figure 2.**
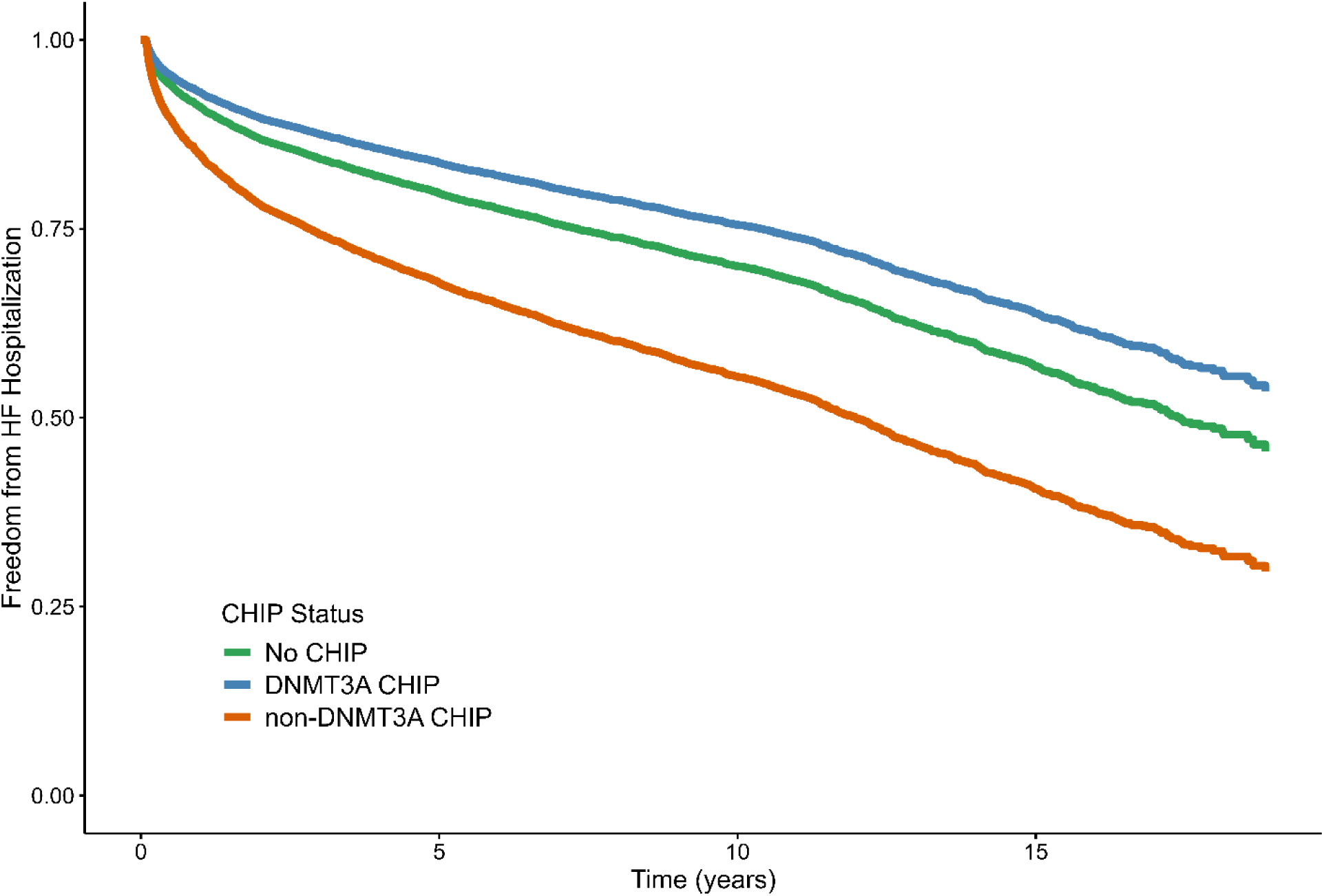
Non-*DNMT3A* and *DNMT3A* CHIP Associate with Incident HF Hospitalization Adjust Kaplan-Meier curves for incident HF Hospitalization. Non-*DNMT3A* CHIP was associated with increased risk of incident HF hospitalization, whereas *DNMT3A* was associated with lower risk. Fine-Gray models are adjusted for age, sex, ancestry, smoking, diabetes, hypertension, hyperlipidemia, body-mass index, prevalent coronary artery disease, HF and atrial fibrillation with overall mortality as a competing risk. HF, heart failure.

After restricting the population to participants without prevalent AF, there were 1398 subjects with incident AF over a median follow-up of 8.13 years (IQR 3.40-11.96 years). There were no significant associations with CHIP or large CHIP for time-to-incident AF diagnosis. Non-*DNMT3A* and *ASXL1* CHIP were associated with time-to-incident AF in univariate models, but only *ASXL1* CHIP remained associated in multivariate models (aHR 2.15 [95% CI 1.15-4.04], p=0.02, **Supplemental Table 7, Supplemental Figure 7**). *ASXL1* CHIP remained associated with time-to-AF diagnosis in sensitivity models truncated at ten years.

## Discussion

For the first time to date in a large, single-center study of 8469 high-risk participants referred for cardiac catheterization, we observed pleiotropic effects of large clones and non-*DNMT3A* CHIP on cardiometabolic traits and CV outcomes (**Central Illustration**). We identified associations of CHIP with obesity, prevalent HF and incident HF hospitalization and AF. Additionally, we recapitulated the known age and mortality associations of CHIP. This study expands the existing CHIP literature on the effect of CHIP across cardiometabolic traits in a comorbid but clinically relevant cohort of high-risk participants referred for cardiac catheterization. Importantly, we show that though *DNMT3A* is the most frequently mutated CHIP gene, variants in non-*DNMT3A* genes drive cardiovascular risk associated with CHIP.

Although we expected to find a greater CHIP prevalence in this cohort enriched for CVD, we found a similar prevalence (5.0%) to previously published population-based CHIP cohorts^6,7,9,27^, but lower than in a recent analysis of five TIMI trials (8.2%) with high baseline CVD^28^. CHIP prevalence in our study was lower than in recent work with a prevalence of 18.4% in 1142 individuals referred for cardiac catheterization at Vanderbilt University Medical Center, however Heimlich et al. used a targeted sequencing approach, with higher sensitivity than the WES used here^29^. With respect to the association between CHIP and demographic characteristics, we found higher prevalence of CHIP in female participants in our cohort; this is discordant with the initial 2014 work by Jaiswal et al., where men had a higher odds of carrying a CHIP mutation^3^. However, our findings are similar to data from other cohorts showing that the effect on sex depends on the driver gene, where there is an increased prevalence of *DNMT3A* mutations in females, but other CHIP driver genes including *TET2* and *ASXL1* are more prevalent in males. We replicated similar findings as have been published between smoking history and *ASXL1* CHIP, but there was no difference for smoking history and overall CHIP^6,7,30,31^.

Interestingly, with respect to cardiometabolic risk factors, we observed an inverse association between CHIP, large CHIP and obesity. To date, there have been conflicting findings between CHIP and obesity. For example, Jaiswal et al. found an inverse relationship between CHIP and BMI^3^, however in 8709 postmenopausal women from the Women’s Health Initiative (WHI), normal and overweight BMI participants had a lower odds of *DNMT3A* and *TET2* CHIP than obese participants^32^. In 47,466 UK Biobank participants, CHIP was associated with higher waist-to-hip-ratio, though only *TET2* CHIP was associated with high BMI^33^. Though we have described for the first time the association of large CHIP clones and obesity, we observed no significant associations for gene-specific analyses and obesity. Conversely, obesity may exacerbate CHIP. *TET2* and *DNMT3A* murine CH models crossed with genetic obesity murine models have shown expansion of CHIP clones and pre-leukemic stem and progenitor cells with heightened levels of pro-inflammatory cytokines^33^. In our cohort there was a strong inverse relationship between age and BMI; this may reflect sarcopenia and frailty. There may be a complex and potentially parabolic relationship between CHIP and BMI, with higher prevalence of CHIP at both extremes of BMI. Future studies should further explore gene-specific metabolic underpinnings of the obesity paradox seen with CHIP in our present work and consider the role of CHIP in organ-specific adiposity.

Interestingly, we did not identify significant associations for CHIP or large CHIP with prevalent CAD. The lack of association may be related to survival bias, the high prevalence of comorbidities and CAD in the CATHGEN cohort, including with the highly sensitive detection of CAD with invasive assessment of coronary lesions in the majority of patients studied. In a recent analysis of five TIMI trials, there was similar baseline prevalence of atherosclerotic CVD between those with and without CHIP and, similar to our observation here, a lower percentage with prior MI in those with CHIP^28^ Taken together, these findings support CHIP as a stronger risk marker for incident disease than prevalent cardiometabolic traits.

To the first time to our knowledge, we identified higher odds of prevalent HF in CHIP carriers in the CATHGEN cohort, with strong associations for non-*DNMT3A* variants. CHIP prevalence was greatest in those with HFrEF, while HFpEF prevalence was intermediate to those without HF. Here we provide key associations for CHIP and prevalent HF, even after adjustment for CAD, adding further support to the growing literature of pathophysiologic mechanisms linking CHIP and HF independent of atherosclerosis. In recent work using ultradeep error-corrected sequencing, prevalence of *TET2* CH (VAF>0.5%) was enriched in HFpEF cases compared to controls^15^. Concordant with prior studies, we also identified greater risk for incident HF hospitalization. Although not significant for overall CHIP, we found associations for non-*DNMT3A* CHIP and incident HF hospitalization. Interestingly, *DNMT3A* CHIP was associated with lower risk of incident HF hospitalization. In a meta-analysis of over 56,000 participants, CHIP was associated with incident HF, specifically *ASXL1*, *TET2* and *JAK2* variants, but the authors did not find significant associations with *DNMT3A* CHIP^13^. More recently, *TET2* CHIP, but not overall CHIP was associated with incident HFpEF-specific hospitalization in 8090 participants from the Jackson Health Study and WHI^14^. In murine models, *TET2* CH exacerbated features of HFpEF compared to wild-type bone marrow. Taken together, these findings suggest that variants in CHIP genes other than *DNMT3A* drive both prevalent and incident HF risk and ongoing work is needed to decipher gene and variant specific risk in clinical and pre-clinical research. Given the complex systemic and metabolic changes in HFpEF, the role of CHIP and its associated inflammatory should be assessed more deeply in future work.

Only large and non-*DNMT3A* CHIP were associated with overall mortality in CATHGEN after adjustment for relevant clinical covariates, however there was no improvement in risk prediction when added to the GRACE score^25^. Unlike prior work, CHIP was not associated with time-to-incident MI or CV death in competing risk regression models in CATHGEN. Large CHIP and non-*DNMT3A*, including *ASXL1* were significant in univariate models though the relationship was attenuated in multivariate models. Interestingly, large CHIP was not associated with HF hospitalization even in univariate models, suggesting risk associated with clone size may be more specific for atherosclerotic phenotypes. These findings are in contrast to data from 424,651 UK Biobank participants, in whom CHIP was associated with incident CVD events (composite MI, CAD or revascularization, stroke or death) with strong associations for large CHIP variants in each gene assessed, including *ASXL1*^34^. In 13,129 individuals in the UK Biobank with established CVD, CHIP and large CHIP were associated with a primary outcome of CVD events, with *TET2* and spliceosome CHIP (*SF3B1*, *SRSF2*, *U2AF1*) having the strongest association with adverse outcomes^35^. In CATHGEN, *ASXL1* CHIP was associated with incident MI or CV death, as well as HF hospitalization in univariate, but not multivariate models. In 235 participants with established MDS, *ASXL1* variants were highly prevalent (40%) and associated with risk of vascular events^36^. The work presented here supports the need for ongoing mutation-specific analyses of CHIP-risk phenotypes and underlying mechanisms, particularly for non-*DNTM3A* CHIP.

We did not identify any significant findings between CHIP and prevalent AF risk, including when looking at large CHIP clones. Concordant with prior work, *ASXL1* CHIP was associated with risk of incident AF. A phenome-wide association study (PheWAS) found baseline CHIP to be associated with incident AF in the UK Biobank^8^. More recently, large *TET2* and *ASXL1* CHIP clones, but not any CHIP or *DNMT3A* CHIP were associated with incident AF in nearly 200,000 participants from the UK Biobank and Atherosclerosis Risk in Communities (ARIC) studies^17^. Further murine models of AF and *TET2* CHIP suggest the therapeutic potential of NLPR3 inflammasome blockade in arrhythmia^18^. Future studies should continue to explore the connection between arrhythmias, CHIP and dysregulated inflammation.

Despite multiple strengths, including long duration of follow-up in a comorbid cohort of diverse participants with ability for detailed EHR review at the individual-participant level, this work has limitations. First, as all participants had at least concern for baseline CVD prompting catheterization referral, the study sample from a cardiac catheterization biorepository limits the generalizability to a broader patient population. As a tertiary referral center, not all participants were followed in Duke University Health system long term. This limited incident data from ICD codes is linked to inpatient and emergency room encounters. We recognize the study may suffer from survival bias for those individuals who may have had CHIP and survived to get catheterization. Likely secondary to limited coverages of the exons of interest in this region, the lower than expected number of *JAK2* V617F variants in our WES data restricted the ability to analyze for *JAK2*-specific associations in CATHGEN.

In summary, in a cohort of participants referred for cardiac catheterization, CHIP and large CHIP were inversely associated with obesity. CHIP and non-*DNMT3A* CHIP were associated with prevalent HF, even in patients with a history of malignancy or chemotherapy exposure. We also identified risk of mortality and incident outcomes for large, non-*DNMT3A* and *ASXL1* CHIP. Further work is warranted to understand the mechanistic underpinnings of the relationships of specific CHIP mutations, inflammation and cardiometabolic disease. Ultimately, utilization and refinement of CHIP as a CVD biomarker may allow for improved precision medicine approaches to patient care.

## Data Availability

All data produced in the present study are available upon reasonable request to the authors

## Funding Acknowledgements

J.A.R. is supported by a grant from the National Heart, Lung and Blood Institute (1R38HL143612) and the Edna and Fred L. Mandel Jr. Foundation.

## Author Contributions

J.A.R., L.C.K and S.H.S. designed the project and statistical analysis plan. W.E.K provided expertise on the CATHGEN biorepository and clinical data, reviewed and edited the manuscript. N.A.N. designed statistical analysis plan for testing associations with atrial fibrillation. A.G.B., P.N. and S.J. provided expertise on CHIP calling methods and quality control. S.J. provided detailed variant review and hematopathologic expert consultation for identification of putative CHIP variants. J.A.R, L.C.K and N.A.N. performed statistical analyses. All authors contributed to manuscript development and critically reviewed the final manuscript.

## Abbreviations

CHIP: clonal hematopoiesis of indeterminate potential

VAF: variant allele fraction

CATHGEN: Catheterization Genetics

CAD: coronary artery disease

MI: myocardial infarction

CV: cardiovascular

CVD: cardiovascular disease

HF: heart failure

AF: atrial fibrillation

## Central Illustration. CHIP Associates with Prevalent and Incident Cardiometabolic Disease in CATHGEN

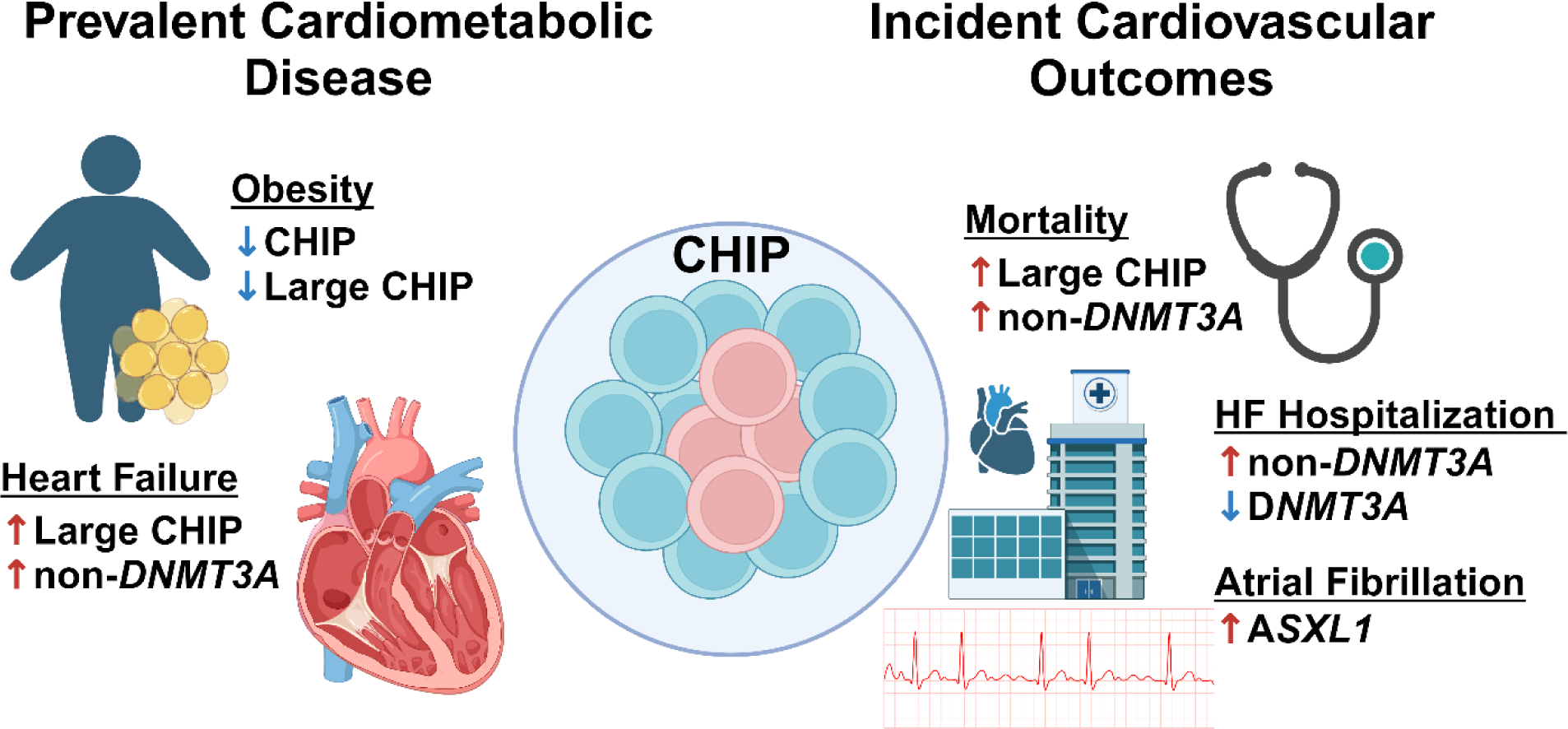

