## Supplemental Methods and Figures for "Clonal Hematopoiesis Associates with Prevalent and Incident Cardiometabolic Disease in High-Risk Individuals"

### SUPPLEMENTAL MATERIAL

#### Supplemental Methods

##### Sample Preparation and Sequencing

Genomic DNA samples were transferred to the Regeneron Genetics Center in 2D matrix tubes (Thermo Scientific), logged into a LIMS (Sapio Sciences), and stored in an automated biobank at -80˚C (LiCONiC TubeStore). Sample quantity was determined by fluorescence (Life Technologies) and quality assessed by running 50ng of sample on a 2% pre-cast agarose gel (Life Technologies). The DNA samples were normalized and 100ng was sheared enzymatically (Kapa Roche) to an average fragment length of 150 base pairs. The sheared genomic DNA was prepared for exome capture with a custom Kapa HyperPlus reagent kit (Kapa Roche) using a fully-automated approach developed at the Regeneron Genetics Center. A unique 6 base pair barcode was added to each DNA fragment during library preparation to facilitate multiplexed exome capture and sequencing. Equal amounts of sample were pooled prior to exome capture with a slightly modified version of IDT's xGen probes; supplemental probes were added to capture regions of the genome well-covered by a previous capture reagent. Captured fragments were bound to streptavidin-conjugated beads and non-specific DNA fragments removed by a series of stringent washes according to the manufacturer's recommended protocol (IDT). The captured DNA was PCR amplified and quantified by qRT-PCR (Kapa Biosystems). The multiplexed samples were sequenced using 75 bp paired-end sequencing on an Illumina v4 HiSeq 2500.

##### Whole Exome Sequencing: Sequence alignment and quality control

Upon completion of sequencing, raw data from each Illumina Hiseq 2500 run was gathered in local buffer storage and uploaded to the DNAnexus platform[1] for automated analysis. Sample-level read files were generated with CASAVA (Illumina Inc., San Diego, CA) and aligned to GRCh38 with BWA-mem[2]. Following completion of cohort sequencing, samples showing disagreement between genetically determined and reported sex, high rates of heterozygosity, low sequence coverage (less than 75% of targeted bases achieving 20X coverage), unusually high degrees of cryptic relatedness, or genetically-identified sample duplicates, were excluded.

##### Somatic Variant Calling and CHIP filtering

The resultant BAM files were processed using the Genome Analysis Toolkit (GATK) v4.1.4.0 Mutect2 v2.7 pipeline.[3-5] A panel of normal (PON) was created from 40 young, healthy CATHGEN participants to help eliminate common sequencing artifacts. Functional annotation was performed with the Funcotator and a single-sample VCF file identifying both SNVs and indels as compared to the reference was output. The VCF files were then filtered to regions of interest based on transcript IDs (**Supplemental Table 2**). Custom R scripts were used to parse each filtered VCF file to identify specific missense variants of interest and frameshift or nonsense (loss-of-function) and splice site variants in relevant genes. For missense variants in specific genes (*CBL*[6]*, CBLB, TET2*[7]), variants were considered somatic if the variant allele frequency (VAF) deviated from the expected distribution of a germline allele by using a binomial test created from the sum of alternate alleles as the number of successes and the sum of the alternate allele count and reference allele count as the number of trials ($p<0.001$). Potential CHIP variants were defined as having a VAF of ≥0.02, with at least four supporting alternate reads, including at least one alternate read in each direction and at least one reference read in each direction. For indels, six or more supporting alternate reads were required. Long indels where either the reference or alternate was greater than six base pairs in length were excluded.

To limit potential inclusion of artifact variants, frameshift variants existing in strings of five or more homopolymer within 10 bases of the variant location were excluded, unless there were a high number of supporting reads (VAF >0.08 and ≥10 alternate reads). Except for *DNMT3A* variants, variants in the first or last 10% of the open reading frame were excluded. This list of potential CHIP variants then underwent manual curation by expert hematopathologist review to further define putative CHIP variants and exclude sequencing artifacts.

##### Clinical Outcomes

A composite outcome of time to first MI or CV death was created. For patients with multiple events of interest, patients were ascertained by their earliest event. For analyses of CV death, 1232 participants had an unknown cause of death and were excluded from analyses. Incident HF hospitalizations were assessed starting 30 days after index catheterization to limit hospitalizations potential related to complications from acute MI. Incident atrial fibrillation analyses were ascertained 6 months after index catheterization to limit overlap with undiagnosed prevalent atrial fibrillation.

##### GRACE Score

The GRACE Score was calculated using the nomogram as detailed under 8. Fox Model for Death between Hospital Admissions and 6 months later (https://www.outcomes-umassmed.org/grace/files/GRACE_RiskModel_Coefficients.pdf). Median imputation was used for missing variables (pulse [0.4% missing], systolic blood pressure [1.9% missing], creatinine [3.9% missing]). Whether or not MI occurred during the current encounter, determined by the enrolling physician, was used as surrogate for abnormal cardiac enzymes. 224 participants (2.6%) had missing data and were set to zero for absent positive enzymes. Killip class was determined by the enrolling physician. 231 participants (2.7%) had missing data for Killip class and were set to Killip Class 1 for No clinical signs of HF. No participants were documented as having cardiac arrest or ST segment deviation on EKG at presentation.

#### Supplemental Figures

##### Supplemental Figure 1. Gene Breakdown and Violin Plot of Variant Allele Fraction for Top 8 CHIP Genes

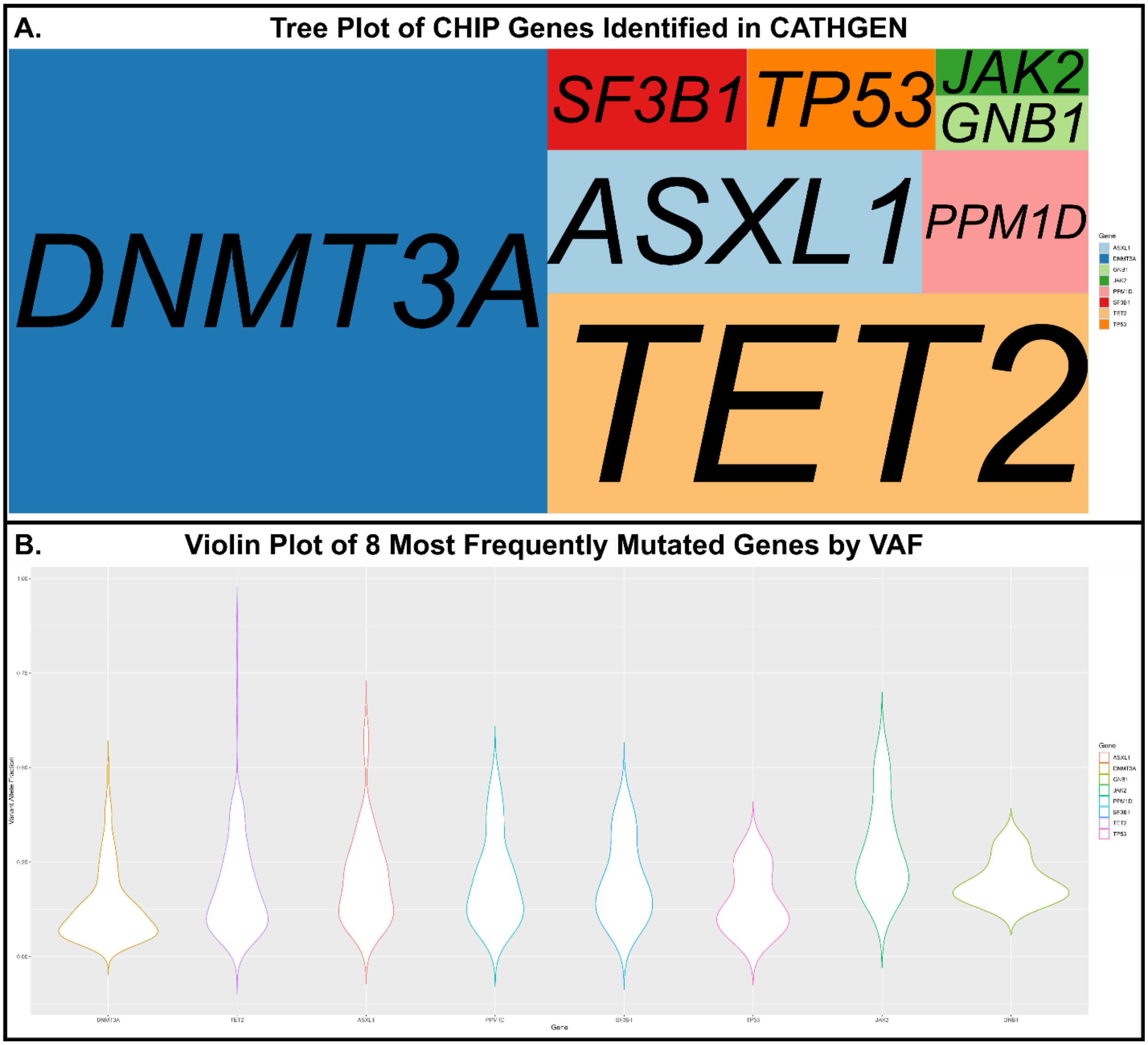


A. Tree plot of top 8 most frequently mutated genes out of 463 total CHIP mutations (*DNMT3A* n=210, *TET2* n=100, *ASXL1* n=45, *PPM1D* n=20, *SF3B1* n=17, *TP53* n=16, *GNB1* n=7, *JAK2* n=6). B. Violin plot by Variant Allele Fraction (VAF) of top eight most frequently mutated genes in CATHGEN.

**Supplemental Figure 2. Age Associated with CHIP in CATHGEN**

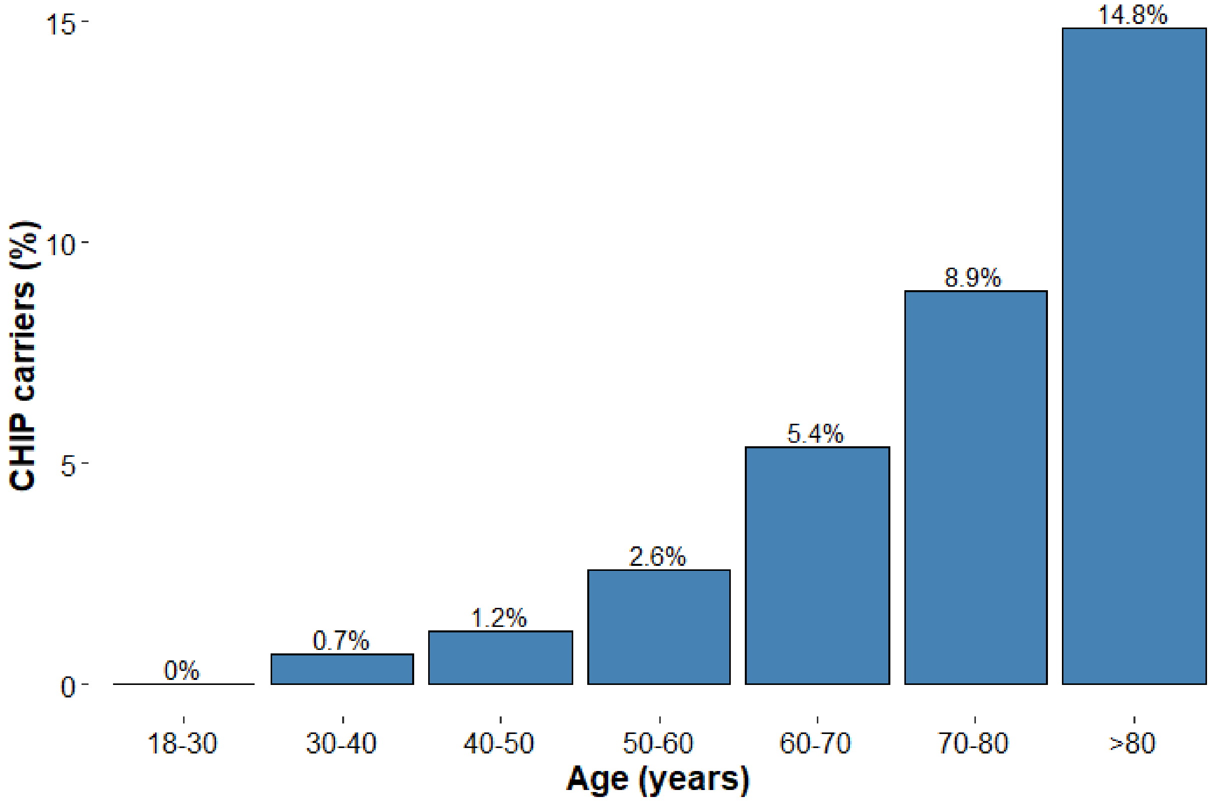


Participants with CHIP were older on average 8.7 years older than those without CHIP (mean 69.5±10.3 years vs 60.8±12.0 years, $p<2x10^{-16}$). There was a higher percentage of CHIP carriers by decade. The overall prevalence of CHIP in the cohort was 5.0%.

**Supplemental Figure 3. Co-mutation plot for CATHGEN participants with more than one CHIP variant**

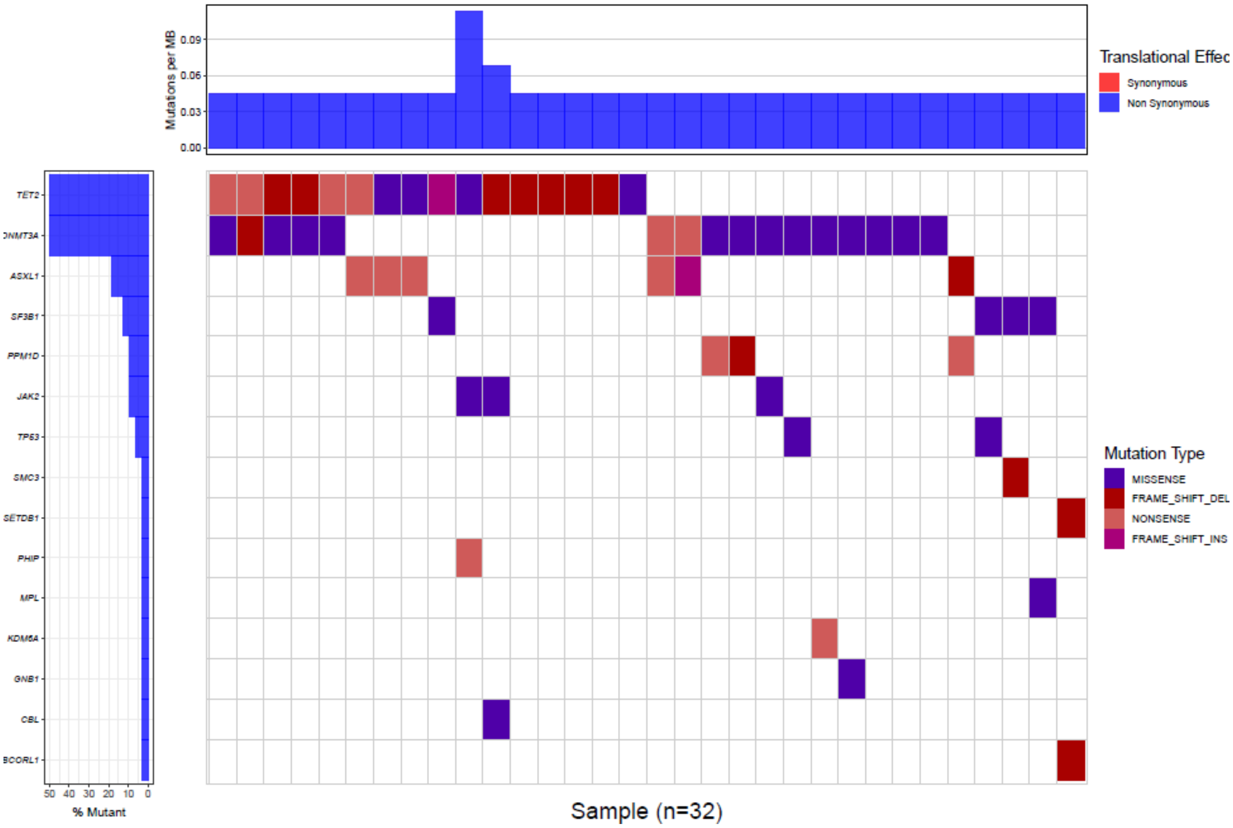


Waterfall plot of CATHGEN participants with more than one CHIP variant identified. *TET2* variants were most frequently identified in participants with more than one CHIP variant. Clones in individuals with more than one CHIP variant were larger (VAF 20.4%±13.3%) than clones in individuals with one CHIP variant (VAF 15.8%±11.5%).

**Supplemental Figure 4. Forest Plot for Association of CHIP and Obesity**

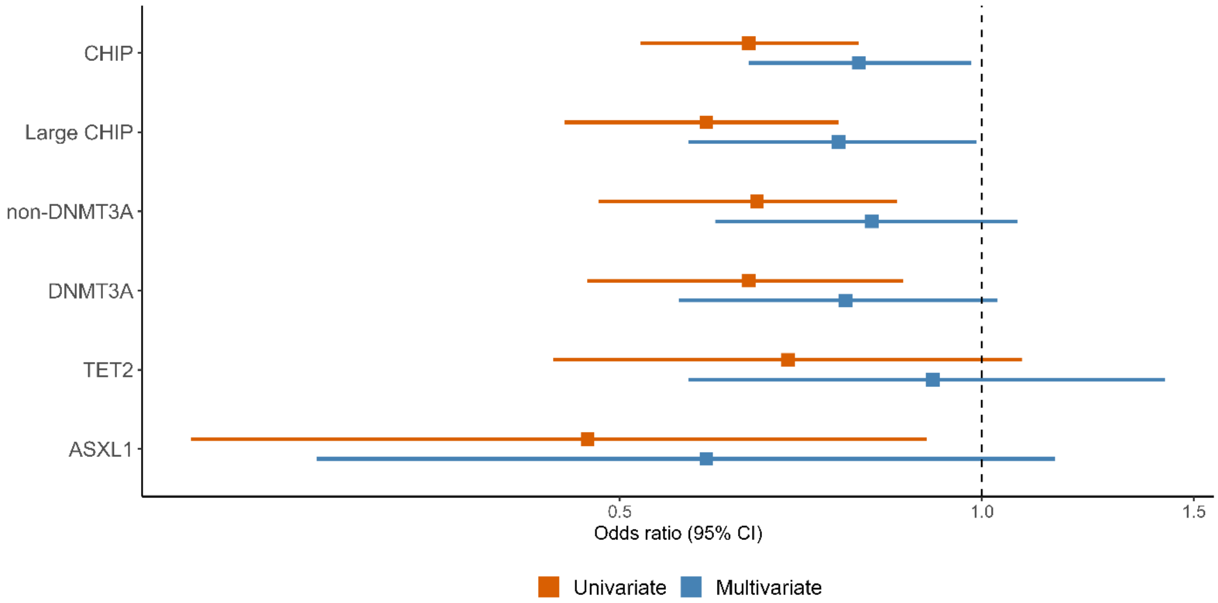


CHIP and large CHIP were inversely associated with obesity in both univariate and multivariate models adjusted for age, sex, ancestry and history of smoking. Non-*DNMT3A*, *DNMT3A* and *ASXL1* CHIP were associated with lower odds of obesity in univariate but not multivariate models.

**Supplemental Figure 5. Prevalence of CHIP by HF Phenotype**

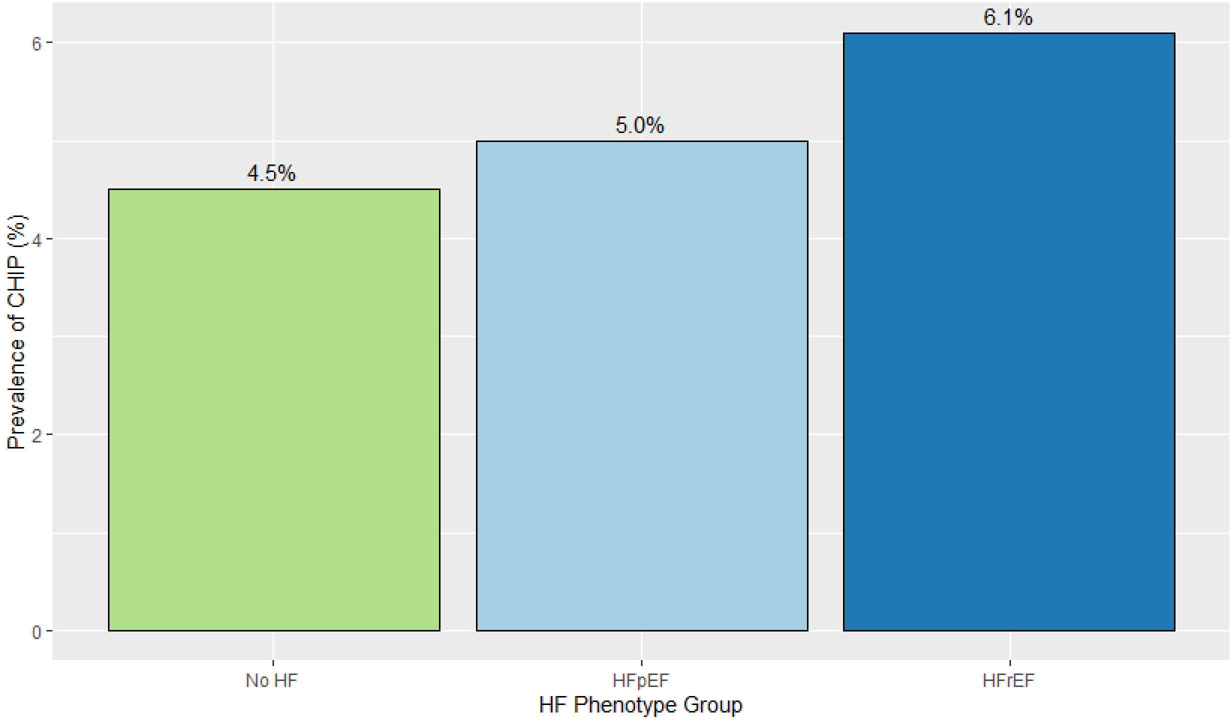


Sensitivity analyses tested the association between heart failure (HF) phenotypes and CHIP. No HF (N=4531, 4.5% CHIP); heart failure with preserved ejection fraction, HFpEF (EF≥50%, N=726, 5.0% CHIP); heart failure with reduced ejection fraction, HFrEF (EF<50%, N=928, 6.1% CHIP). ANOVA p=0.04, chi-squared p=0.04 for HFrEF vs. No HF.

**Supplemental Figure 6. Large CHIP and non-*DNMT3A* CHIP associate with time-to-overall mortality in CATHGEN**

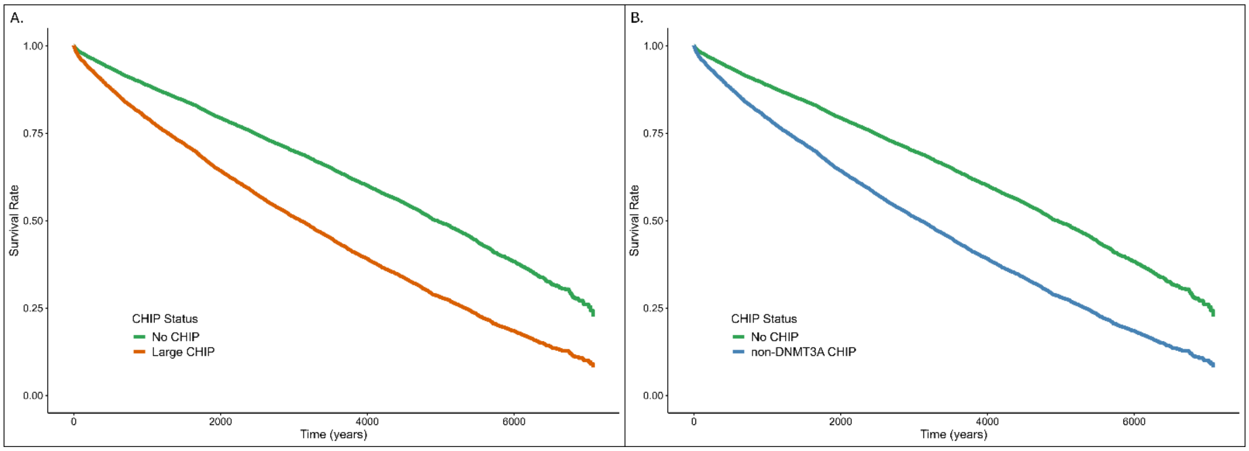


Adjusted Kaplan-Meier curves for the association of large CHIP clones (VAF≥10%, Panel A). and non-*DNMT3A* CHIP variants (Panel B). Multivariate cox models are adjusted for age, sex, ancestry, smoking, diabetes, hypertension, hyperlipidemia, body-mass index, prevalent coronary artery disease, heart failure and atrial fibrillation.

**Supplemental Figure 7. *ASXL* CHIP Associates with Risk of Incident Atrial Fibrillation in CATHGEN**

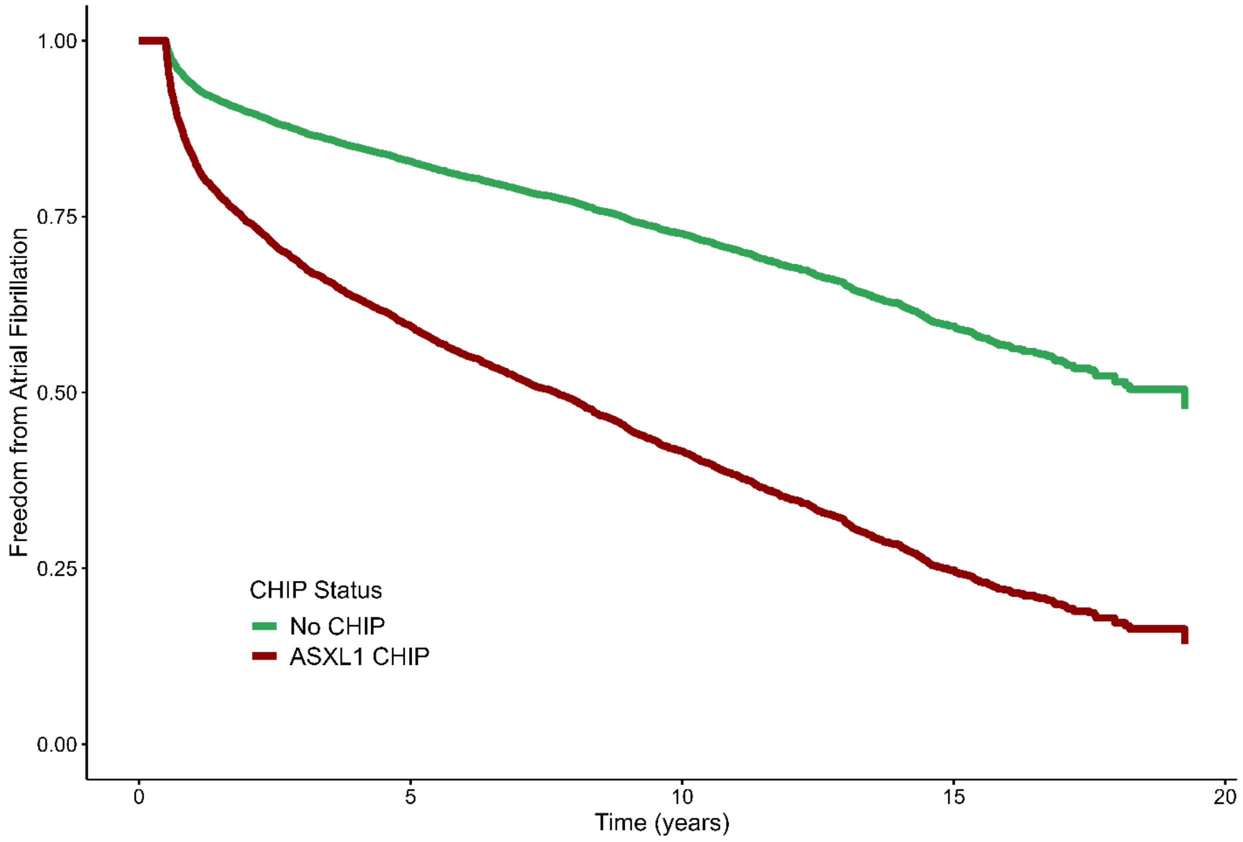


Adjusted Kaplan-Meier curve for *ASXL1* CHIP and incident atrial fibrillation. Models are adjusted for age, sex, ancestry, smoking, diabetes, hypertension, hyperlipidemia, body-mass index, prevalent coronary disease and heart failure.
